# The COVID-19 Spread Patterns in Italy and India: A Comparison of the Current Situations

**DOI:** 10.1101/2020.06.21.20136630

**Authors:** Hemanta K. Barauh

## Abstract

Epidemiological mathematical models and time series models can be used to forecast about the spread of an infectious disease. In this article, without using such models, we are going to show how exactly the pattern evolves day by day once a pattern is seen to be approximately followed by the data. Although in Italy as well as in India the novel corona virus appeared on the same day, in Italy the spread is nearly logarithmic by now and in India it is nearly exponential even now.

## Introduction

Various epidemiological models to study the spread of infectious diseases are available in the literature. Meyers [1] studied the use of the Susceptible-Infected-Recovered (SIR) model for infectious diseases affecting a small region. Kamp [2] concluded that the transmission network strongly influences speed and range of spread of epidemics. Liu *et. al*. [3] studied theoretically the Susceptible-Infectious-Susceptible (SIS) pandemic model using time delays in the thresholds. Majeed and Shawka [4] studied mathematically the SIS model of epidemic growth. Wu *et. al*. [5] used the Susceptible-Exposed-Infectious-Recovered (SEIR) model in a simulation study of the COVID-19 spread. Recently Grant [6] has shown that SEIR model does not work properly in the case of the spread of COVID-19. There are certain other models too such as the Susceptible-Infectious-Recovered-Dead (SIRD) model [7]. Indeed, quite a few more epidemiological models are in use to study the spread of infectious diseases. It has been seen that some models do forecast about the spread very nicely while some others have not been very successful.

Time series models using the auto-regressive integrated moving average (ARIMA) method have also been used successfully by a few authors for forecasting the COVID-19 spread in India. Poonia and Azad [8] and Azad and Poonia [9] studied the situation in two phases using the ARIMA method. Their forecasts were close to the actual values observed later. Basu [10] studied time dependent spread of the virus in India using his own model. As per his forecasts expressed in the form of a graph, the total number of cases in India should have crossed 200,000 in the beginning of June, and his forecast has been found to be true.

In this work, we have two objectives. First, we shall compare the current spread patterns in Italy and India. From the graphs of the current data on total number of cases that includes active cases, recovered cases and deaths, some mathematical pattern would be apparent. We would find out the current approximate patterns of the spread in these two countries. Secondly, we would like to show how the patterns evolve day by day. Hypothesizing about a particular mathematical model as the underlying spread pattern is one thing, while studying the changes day by day looking into the recent data is quite another. We would show how the logarithmic function is being followed by the total number of cases in Italy, and how in India it is following a nearly exponential function, while the patterns are changing slowly and steadily.

We would like to state that while trying to find a mathematical model regarding the spread pattern of a pandemic, the epidemiological models assume that in the first phase of the spread the mathematical pattern is exponential. In fact, this assumption is a very simplified one. Initially, the pattern would be nonlinear but assuming it to be exponential right from the beginning is not actually valid. Indeed, it can in reality be only approximately exponential, because if the pattern is exponential, it would only mean that the pattern would remain so until everything is finished. When the pandemic happens to retard, it is said that the curve is getting flattened. It is important to examine following what pattern the growth starts retarding. In any phase, we cannot say that a particular growth pattern is being strictly followed. When the pandemic would finally come to an end, it would mean that the pattern is a straight line parallel to the time axis. Before that happens, the pattern may be approximately logarithmic. Indeed a logarithmic growth is also a growth, however slow it is.

We are in this article going to discuss and compare the current growth patterns of COVID-19 in Italy and India. We would show that in Italy the current growth is approximately logarithmic while in India it is still growing approximately exponentially, although as per data published by Worldomters.info [11], on February 15, 2020, both in Italy and in India the total numbers of cases were equal to 3.

## Methodology

Let a function *f*(*x*) be logarithmic in *x*. Then

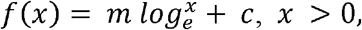

where *m* and *c* are constants. If we need to fit such a curve from some observed data, we have to estimate the *parameters* from the data. That would need a method such as the method of least squares to estimate the two parameters. However, in the case that we are currently going to discuss, we would assume that from a certain value of *x* the concerned curve is logarithmic. Therefore taking that starting value of *x* as 1, if we want to verify whether the curve is approximately logarithmic, then the value of the constant *c* is already known to us, and therefore in that kind of a situation we would need to estimate only the value of *m*. If we now see that the estimated values of *m* found from

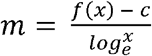

for a few consecutive values of *x* are very nearly constant, then it would assert that the underlying function *f*(*x*) is nearly logarithmic.

Let a function *g*(*x*) be exponential in *x*. Then

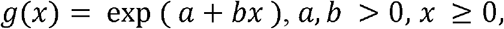

where *a* and *b* are constants. Then

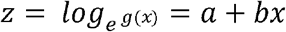

would be linear in *x*. Although there are two parameters involved, when we proceed to apply this model in the exponential phase of a pandemic, we can take some particular date as the base to describe a few dates thereafter with reference to the base date. Accordingly, the value of the constant *a* would be available to us already and we would have to find an estimate of the parameter *b*. When we would observe that the estimated values of *b* are very nearly constant, we would be able to say that the pattern is approximately exponential.

### The Italian Situation

As per the Worldometers.info data [11], on February 15 there were 3 cases reported in Italy. From the graphs it can be seen that by the middle of March the spread pattern became highly nonlinear. By the end of April, the process of curve flattening had started. In other words, the pattern changed by the end of April from an approximately exponential form to a logarithmic form. We are explaining the matters in a rather simple way. But it can actually be checked whether it was really so. In Table-1, we are showing the values of the estimated values of *m* from June 10 to June16 for Italy. We shall use here the relation

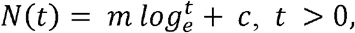

where *c* = 205463, the value of *N*(*t*) on April 30, 2020. Here *t* = 1 on the base date, April 30.

**Table-1:** The values of *m* in Italy from June 10 to June 16.

| Dates | $t$ | $N(t)$ | $N(t) - c$ | $m$ |
| --- | --- | --- | --- | --- |
| June 10 | 42 | 235763 | 30300 | 8106.65 |
| June 11 | 43 | 236142 | 30679 | 8156.70 |
| June 12 | 44 | 236305 | 30842 | 8150.22 |
| June 13 | 45 | 236651 | 31188 | 8193.00 |
| June 14 | 46 | 236989 | 31526 | 8234.25 |
| June 15 | 47 | 237290 | 31827 | 8266.43 |
| June 16 | 48 | 237500 | 32037 | 8275.72 |

The date could have been taken as any other date nearby the approximate turning point from nearly exponential to a hypothesized nearly logarithmic spread pattern.

It can be seen that the values of *m* are very nearly constant, but they are still increasing although very slowly. Therefore if we use the average of these values of *m* to extrapolate for June 17, we would actually end up underestimating the value of *N*(*t*) for *t =* 49. Taking the value of *m* with June 16 as the base date and with *m* = 8275.72 as on June 16, we get

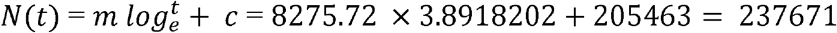

while the actual observed value of the total number of cases in Italy on June 17 was 237828. In other words, in Italy, the spread pattern is indeed approximately logarithmic at least since April 30, the date that we have taken as the approximate turning point. The pattern would continue to be approximately logarithmic until the pandemic comes to a stop after which the pattern would be a straight line parallel to the time axis.

### The Indian Situation

It is apparent from the graph published by Worldometers.info [11] that in India the spread pattern is still approximately exponential. It was seen that the function

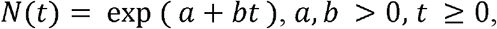

fits the data of spread in India [12] approximately.

To estimate the value of the parameter *b* at some point of time we would need data about the total number of cases for a few days prior to that. If the values of Δz, the first order differences of *z* are seen to be nearly constant, then we can say that the pattern is nearly exponential. It was observed in [12] that the values of Δz have been following a reducing trend with some irregularities in between which is inherent in the case of a time series of this type of a pandemic. In Table-2, we have shown the values of *N*(*t*) and Δz for 3 days, June 8 to June 10.

**Table-2:** Estimation of *b* for India from 3 consecutive days.

| Dates | $N(t)$ | $Z$ | $\Delta z$ |
| --- | --- | --- | --- |
| June 10 | 287155 | 12.567777 | 0.04405 |
| June 9 | 274780 | 12.523726 | 0.03274 |
| June 8 | 265928 | 12.490980 | 0.03226 |

The average of these 3 values of Δz is 0.03635. Taking 0.036 as an estimate of the parameter *b* and taking June 10 as the base date the following expected values of the total number of cases in India from June 11 to June 17 can be computed.

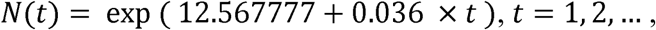

for *t* = 1, 2, …, we would get the expected values for June 11, 12, and so on.

We are showing in Table-3 the expected values of *N*(*t*) from June 11 to June 17 for India. It can be seen that the differences between the observed values and the expected values are negligibly small with reference to the largeness of the observed values.

**Table-3:** Comparison of Observed and Expected Values of the Total Cases in India.

| Dates | Observed Values | Expected Values |
| --- | --- | --- |
| June 11 | 298283 | 297681 |
| June 12 | 309603 | 308593 |
| June 13 | 321626 | 319904 |
| June 14 | 332783 | 331630 |
| June 15 | 343026 | 343786 |
| June 16 | 354161 | 356388 |
| June 17 | 367264 | 369452 |

This shows that at least up to June 17 the pattern of spread in India is nearly exponential. Our objective was not to forecast, but to show that the nearly exponential pattern is still continuing in India. Indeed, our method is valid for short term forecasting only. As has been mentioned earlier, the values of Δz were seen to be in a reducing trend. So this method is unsuitable for long term forecasting.

## Conclusions

Although the pandemic had started in Italy and India on the same date, February 15, 2020, the situations soon became very different in the two countries. In Italy, the nearly exponential pattern could be observed before the middle of April, and currently it is increasing logarithmically. In India however, in April the spread was just highly nonlinear and currently it is nearly exponential. This shows that a study regarding the total number of cases in the world as a whole cannot follow one single mathematical model, because whereas in India the spread pattern is continuing to be nearly exponential, in Italy it is the inverse function – the logarithmic function – being followed by the data.

## Data Availability

The data used are from Worldometers.info.

https://www.worldometers.info/coronavirus/coronavirus-cases/

